# Mendelian randomization analysis identified genes pleiotropically associated with central corneal thickness

**DOI:** 10.1101/2021.02.27.21252574

**Authors:** Zhikun Yang, Jingyun Yang, Di Liu, Weihong Yu

## Abstract

**Objective:** To prioritize genes that were pleiotropically or potentially causally associated with central corneal thickness (CCT).

**Methods:** We applied the summary data-based Mendelian randomization (SMR) method integrating summarized data of genome-wide association study (GWAS) on CCT and expression quantitative trait loci (eQTL) data to identify genes that were pleiotropically associated with CCT. We performed separate SMR analysis using CAGE eQTL data and GTEx eQTL data. SMR analysis were done for participants of European and East Asian ancestries, separately.

**Results:** We identified multiple genes showing pleiotropic association with CCT in the participants of European ancestry. *CLIC3* (ILMN_1796423; P_SMR_=4.15×10^−12^), *PTGDS* (ILMN_1664464; P_SMR_=6.88×10^−9^) and *C9orf142* (ILMN_1761138; P_SMR_=8.09×10^−9^) were the top three genes using the CAGE eQTL data, and *RP11-458F8*.*4*(ENSG00000273142.1; P_SMR_=5.89×10^−9^), *LCNL1* (ENSG00000214402.6; P_SMR_=5.67×10^−8^), and *PTGDS* (ENSG00000107317.7; P_SMR_=1.92×10^−7^) were the top three genes using the GTEx eQTL data. No genes showed significantly pleiotropic association with CCT in the participants of East Asian ancestry after correction for multiple testing.

**Conclusion:** We identified several genes pleiotropically associated with CCT, some of which represented novel genes influencing CCT. Our findings provided important leads to a better understanding of the genetic factors influencing CCT, and revealed potential therapeutic targets for the treatment of primary open-angle glaucoma and keratoconus.

## Introduction

The cornea is a highly collagenous and transparent tissue through which light reaches the interior structures of the eye. Previous studies highlighted the importance of central corneal thickness (CCT) in relation to several ocular and non-ocular conditions. For example, decrease in CCT is significantly associated with intraocular pressure (IOP)[1]. Thinner CCT has been demonstrated as an important feature of keratoconus and a risk factor for primary open-angle glaucoma (POAG) in patients with ocular hypertension[2-7]. Keratoconus is the leading cause of corneal transplants worldwide, with an estimated prevalence of 0.14%[8], while POAG is the most common cause of irreversible blindness worldwide, accounting for approximately 70% of all the cases of glaucoma[9].

Epidemiologic studies have shown that CCT differs among ethnic groups, with Europeans having higher CCT values than Africans, and Asians showing a larger variation in CCT[10]. CCT is highly heritable, with an estimated heritability ranging from 68% to 95%[11-13]. Previous genome-wide association studies (GWAS) identified a number of CCT-associated loci in Europeans and Asians, such as genetic loci in *ZNF469, FOXO1, LRRK1* and *IBTK*[14-20]. Recent genetic studies revealed additional novel loci associated with CCT, some of which conferred relatively high risks of keratoconus and POAG, highlighting the potential involvement of CCT-associated genes underlying the pathogenesis of keratoconus and POAG[21, 22].

Mendelian randomization (MR) uses genetic variants as the proxy to randomization and is a promising tool to explore pleotropic/potentially causal effect of an exposure (e.g., gene expression) on the outcome (e.g., CCT)[23]. MR could reduce confounding and reverse causation that are commonly encountered in traditional association studies, and has been successful in identifying gene expression probes or DNA methylation loci that are pleiotropically/potentially causally associated with various phenotypes, such as neuropathologies of Alzheimer’s disease and severity of COVID-19[24, 25]. In this paper, we applied the summary data-based MR (SMR) method integrating summarized GWAS data for CCT and cis-eQTL (expression quantitative trait loci) data to prioritize genes that are pleiotropically/potentially causally associated with CCT.

## Methods

### Data sources

#### eQTL data

In the SMR analysis, cis-eQTL genetic variants were used as the instrumental variables (IVs) for gene expression. We performed SMR analysis using gene expression in blood due to unavailability of eQTL data for the eye. Specifically, we used the CAGE eQTL summarized data[26], which included 2,765 participants, and the V7 release of the GTEx eQTL summarized data[27], which included 338 participants. The eQTL data can be downloaded at https://cnsgenomics.com/data/SMR/#eQTLsummarydata.

#### GWAS data for corneal thickness

The GWAS summarized data for CCT were provided by a recent genome-wide association meta-analysis of CCT[21]. The results were based on meta-analyses of 1000 genomes phase 1 imputed GWASs on CCT, with a total of 19 cohorts from the International Glaucoma Genetics consortium (IGGC). Specifically, the meta-analysis for participants of European ancestry included 14 cohorts with a sample of size of 17,803, and the meta-analysis for participants of Asian ancestry included 5 cohorts with a sample size of 8,107. All participating studies assumed an additive genetic model, adjusting for age, sex and at least the first five principal components. The GWAS summarized data can be downloaded at https://datashare.is.ed.ac.uk/handle/10283/2976.

#### SMR analysis

We conducted the SMR analysis with cis-eQTL as the IV, gene expression as the exposure, and CCT as the outcome. The analysis was done using the method as implemented in the software SMR. Detailed information regarding the SMR method was reported in a previous publication[28]. In brief, SMR applies the principles of MR to jointly analyze GWAS and eQTL summary statistics in order to test for pleotropic association between gene expression and a trait due to a shared and potentially causal variant at a locus. We also conducted the heterogeneity in dependent instruments (HEIDI) test to evaluate the existence of linkage in the observed association. Rejection of the null hypothesis (i.e., P_HEIDI_<0.05) indicates that the observed association could be due to two distinct genetic variants in high linkage disequilibrium with each other. We adopted the default settings in SMR (e.g., P_eQTL_ <5 × 10^−8^, minor allele frequency [MAF] > 0.01, removing SNPs in very strong linkage disequilibrium [LD, r^2^ > 0.9] with the top associated eQTL, and removing SNPs in low LD or not in LD [r^2^ <0.05] with the top associated eQTL), and used false discovery rate (FDR) to adjust for multiple testing.

We used Affymetrix exon array S1.0 platforms to annotate the transcripts. To functionally annotate putative transcripts, we conducted functional enrichment analysis using the functional annotation tool “Metascape” for the top 15 tagged genes[29]. Gene symbols corresponding to top hit genes were used as the input of the gene ontology (GO) and Kyoto Encyclopedia of Genes and Genomes (KEGG) enrichment analysis.

Data cleaning and statistical/bioinformatical analysis was performed using R version 4.0.3 (https://www.r-project.org/), PLINK 1.9 (https://www.cog-genomics.org/plink/1.9/) and SMR (https://cnsgenomics.com/software/smr/).

## Results

### Basic information of the summarized data

The number of participants of the CAGE eQTL data is much larger than that of the GTEx eQTL data, so is the number of eligible probes. The sample size of the GWAS data for the European ancestry is much large than that for the Asian ancestry, so is the number of eligible genetic variants. The detailed information was shown in **Table 1**.

### SMR analysis in the European population

In participants with European ancestry, we identified multiple genes showing pleiotropic association with CCT after correction for multiple testing using FDR (**Table 2**). Specifically, using the CAGE eQTL data, our SMR analysis identified 12 genes that were pleiotropically/potentially causally associated with CCT, with *CLIC3* (ILMN_1796423; P_SMR_=4.15×10^−12^), *PTGDS* (ILMN_1664464; P_SMR_=6.88×10^−9^) and *C9orf142* (ILMN_1761138; P_SMR_=8.09×10^−9^) being the top three genes (**Figure 1**). GO enrichment analysis of biological process and molecular function showed that the significant genes were involved in xxx GO terms. Concept network analysis of the identified genes revealed xxxx (**Figure 2**). More information could be found in **Table S2**.

**Figure 1.**
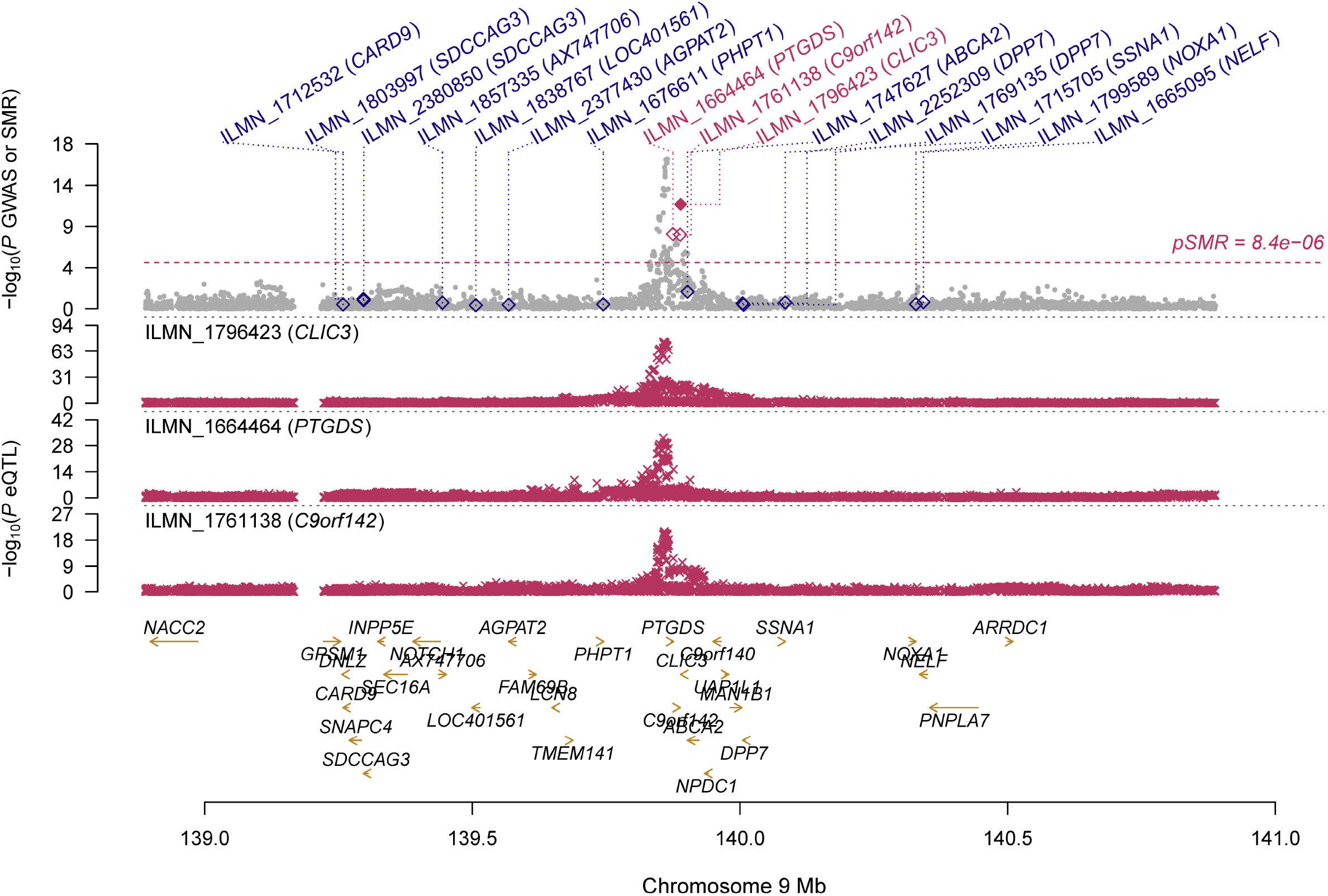
Prioritizing gene around *CLIC3, PTGDS* and *C9orf142* in pleiotropic association with CCT in the participants of European ancestry using CAGE eQTL data. Top plot, grey dots represent the -log_10_(*P* values) for SNPs from the GWAS of CCT, and rhombuses represent the -log_10_(*P* values) for probes from the SMR test with solid rhombuses indicating that the probes pass HEIDI test and hollow rhombuses indicating that the probes do not pass the HEIDI test. Middle plot, eQTL results. Bottom plot, location of genes tagged by the probes. CCT, central corneal thickness; GWAS, genome-wide association studies; SMR, summary data-based Mendelian randomization; HEIDI, heterogeneity in dependent instruments; eQTL, expression quantitative trait loci

**Figure 2.**
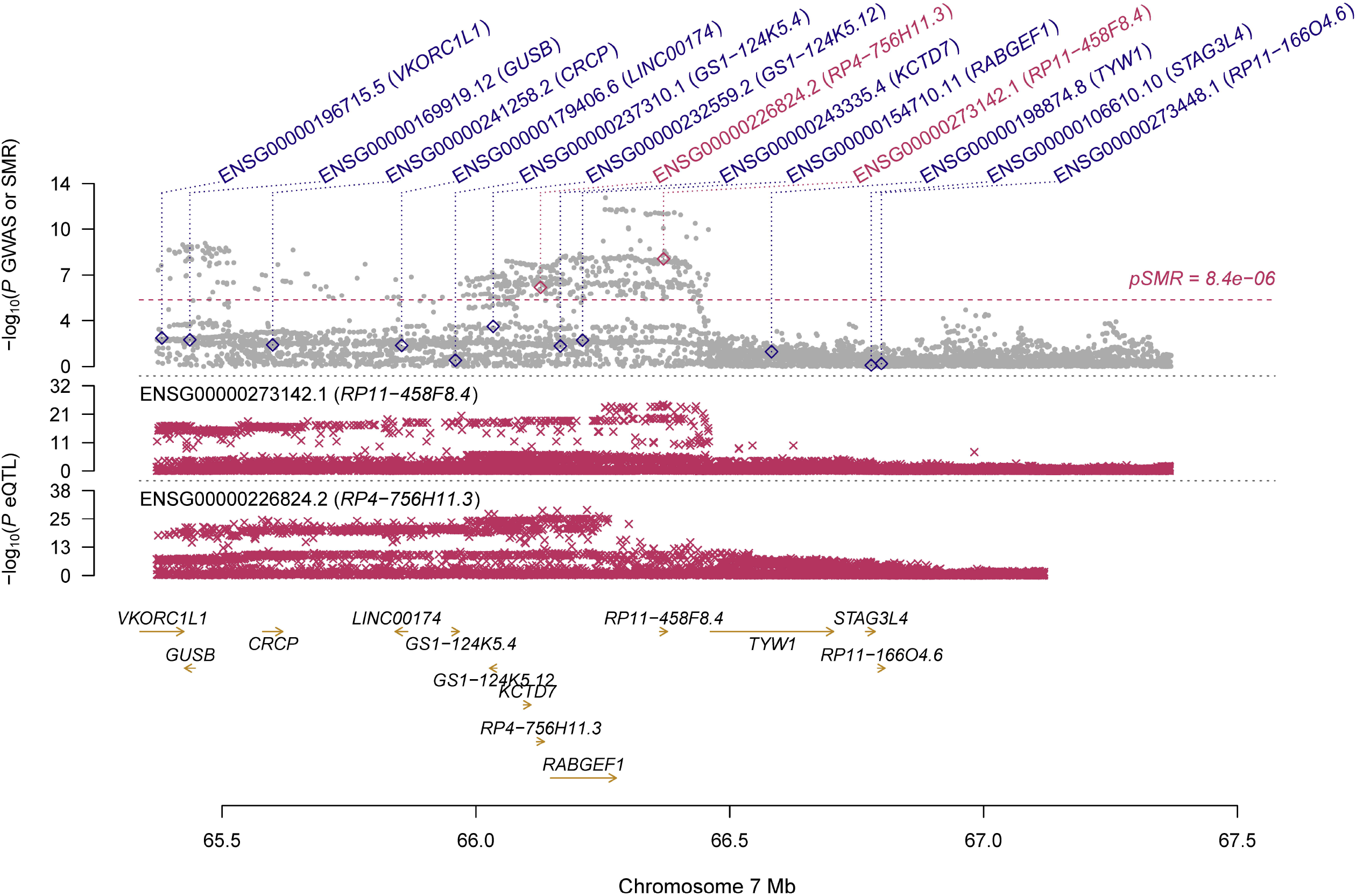
Prioritizing gene around *RP11-458F8*.*4* in pleiotropic association with CCT in the participants of European ancestry using GTEx eQTL data. Top plot, grey dots represent the -log_10_(*P* values) for SNPs from the GWAS of CCT, and rhombuses represent the -log_10_(*P* values) for probes from the SMR test with hollow rhombuses indicating that the probes do not pass the HEIDI test. Middle plot, eQTL results. Bottom plot, location of genes tagged by the probes. CCT, central corneal thickness; GWAS, genome-wide association studies; SMR, summary data-based Mendelian randomization; HEIDI, heterogeneity in dependent instruments; eQTL, expression quantitative trait loci

Using the GTEx eQTL data, our SMR analysis identified six genes that were pleiotropically/potentially causally associated with CCT, with *RP11-458F8*.*4* (ENSG00000273142.1; P_SMR_=5.89×10^−9^), *LCNL1* (ENSG00000214402.6; P_SMR_=5.67×10^−8^), and *PTGDS* (ENSG00000107317.7; P_SMR_=1.92×10^−7^) being the top three genes (**Figure 3** & **4**). GO enrichment analysis of biological process and molecular function showed that the significant genes were involved in xxx GO terms. Concept network analysis of the identified genes revealed xxxx (**Figure 2**). More information could be found in **Table Sx**. It should be noted that the gene *PTGDS* was the top gene showing significant pleiotropic association in both SMR analyses.

**Figure 3.**
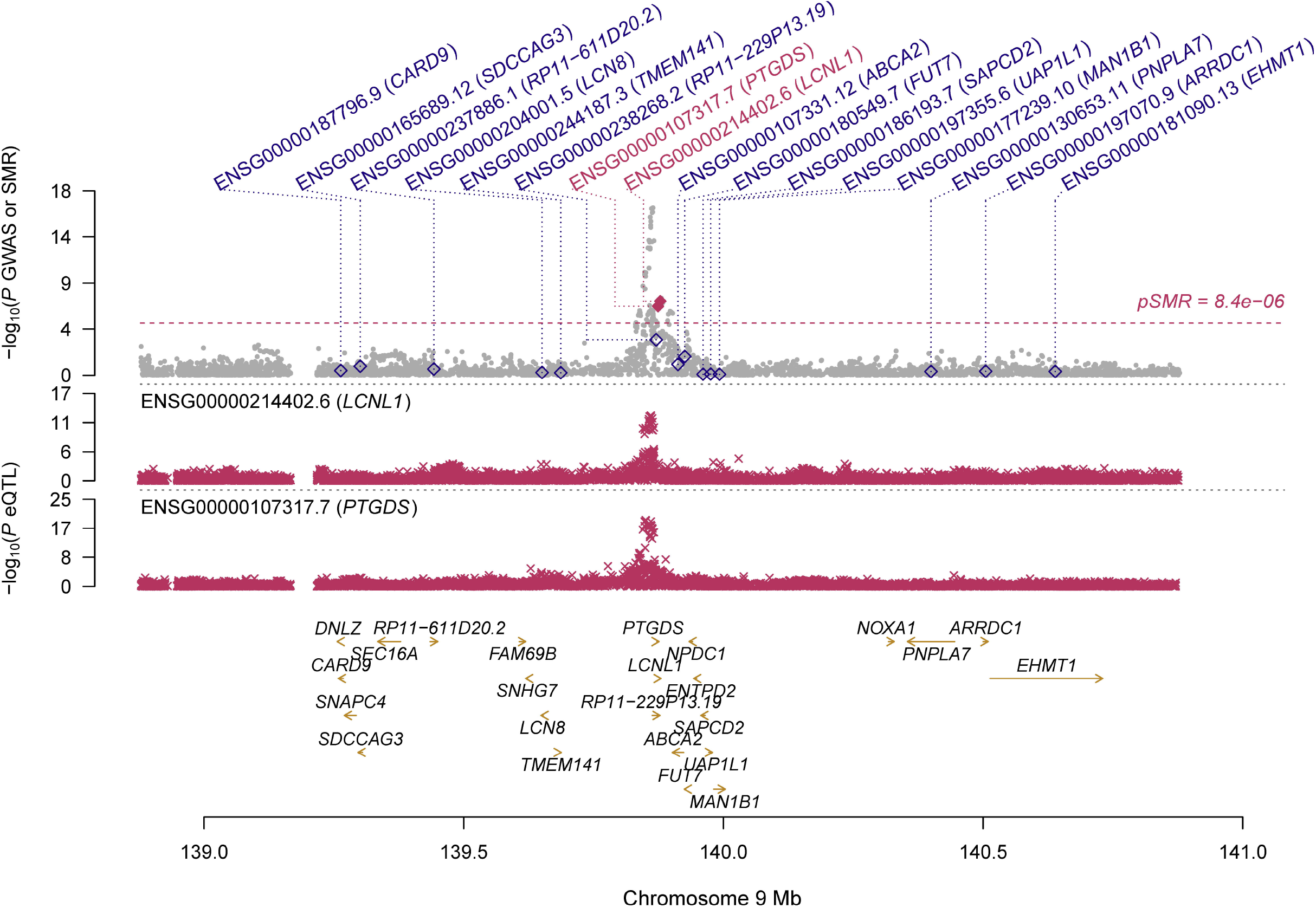
Prioritizing gene around *LCNL1* and *PTGDS* in pleiotropic association with CCT in the participants of European ancestry using CAGE eQTL data. Top plot, grey dots represent the -log_10_(*P* values) for SNPs from the GWAS of CCT, and rhombuses represent the -log_10_(*P* values) for probes from the SMR test with solid rhombuses indicating that the probes pass HEIDI test and hollow rhombuses indicating that the probes do not pass the HEIDI test. Middle plot, eQTL results. Bottom plot, location of genes tagged by the probes. CCT, central corneal thickness; GWAS, genome-wide association studies; SMR, summary data-based Mendelian randomization; HEIDI, heterogeneity in dependent instruments; eQTL, expression quantitative trait loci

### SMR analysis in the East Asian population

In participants with East Asian ancestry, we identified no genes showing significant pleiotropic association with CCT after correction for multiple testing using FDR (**Table 3**). Specifically, using the CAGE eQTL data, we found that two genes, *PASK* (ILMN_1754858, P_SMR_=7.98×10^−5^; ILMN_1667022, P_SMR_=8.24×10^−5^) and *HIATL1* (ILMN_1703229, P_SMR_=4.30×10^−4^; ILMN_1737964, P_SMR_=5.84×10^−4^), each of which was tagged by two probes, were among the top hits in the SMR analysis. However, none of the genes survived multiple comparison. Using the GTEx eQTL data, we found that several genes overlap with the top genes in the SMR analysis using CAGE eQTL data, including *MAP7D1, HIATL1, POLA2* and *ACADM*. Again, none of the genes survived multiple comparison.

## Discussion

In the present study, we integrated summarized data of GWAS on CCT and eQTL data in the MR analysis to explore putative genes that showed pleiotropic/potentially causal association with CCT. In the participants of European ancestry, we identified multiple genes showing significantly pleiotropic association with CCT, some of which represented novel genes associated with CCT. Our findings provided important leads to a better understanding of the genetic factors influencing CCT, and revealed potential therapeutic targets for the treatment of POAG and keratoconus.

We found that *PTGDS* (prostaglandin D2 synthase) showed significantly pleiotropic association with CCT in the participants of European ancestry using both CAGE and GTEx eQTL data. *PTGDS* encodes the glutathione-independent prostaglandin D2 synthase which catalyzes the conversion of prostaglandin H2 (PGH2) to prostaglandin D2 (PGD2), an important marker for keratocytes[30, 31]. A genetic polymorphism in *PTGDS* (rs11145951) was reported to be associated with CCT in the European population (P=9.20×10^−12^) but not in the Asian population (P=2.30×10^−2^)[15]. The association of this polymorphism with CCT did not reach genome-wide significance in the Latino population (P=1.15×10^−5^), suggesting ethnic-specific effect of this genetic polymorphism[32]. *PTGDS* was found to be highly expressed in corneal endothelial cells (CECs)[33], ranked the among the top 50 most highly expressed genes in CECs[34]. When using a novel dual media culture approach for the *in vitro* expansion of primary human corneal endothelial cells (hCECs), the expression of *PTGDS* increased by 12.64 folds following exposure of cultivated hCECs from proliferative (M4) to maintenance (M5) medium[35]. Given that CCT was an important feature for keratoconus and a potential risk factor for POAG, these findings, together with ours, demonstrated the important role of *PTGDS* in influencing CCT and highlighted the potential of this gene as a promising target for the prevention and treatment of keratoconus and POAG. *RP11-458F8*.*4* showed the most significantly pleiotropic association with CCT using GTEx eQTL data in the participants of European ancestry (**Table 2**). *RP11-458F8*.*4* is a long intergenic noncoding RNA (lincRNA). LincRNAs exercise various tissue-specific functions such as remodeling chromatin and genome architecture, RNA stabilization and transcription regulation[36]. *RP11-458F8*.*4* was reported to be associated with various types of malignant tumors. For example, it was upregulated in late stage colon cancers, and its expression may be involved in the progression of colon cancer[37]. It was found to be a prognostic lincRNA in high-grade serous epithelial ovarian cancer[38], and was differently expressed in patients with breast cancer[39]. No studies have reported the association of this gene with CCT. As a result, further investigation is needed to elucidate the exact functions of this gene and examine its role in influencing CCT.

*VKORC1L1* (vitamin K epoxide reductase complex subunit 1 like 1) was pleiotropically associated with CCT in in the participants of European ancestry using CAGE eQTL data. *VKORC1L1* can mediate vitamin K-dependent intracellular antioxidant functions[40]. A genetic variant in *VKORC1L1* (rs11763147) was found to be associated with CCT in a combined meta-analysis of the European and the Asian samples (4.0×10^−9^)[15]. Another genetic variant (rs10563220) located upstream of *VKORC1L1* was found to be significantly associated with intraocular pressure in a GWAS of 8,552 Chinese participants[41]. More studies are needed to elucidate the exact functions of *VKORC1L1* in association with CCT, and whether/how it is involved in the pathogenesis of POAG and keratoconus.

*RUNX2* (runt-related transcription factor 2) showed significantly pleiotropic association with CCT in the participants of European ancestry using the CAGE eQTL data (**Table 2**). *RUNX2* is a member of the RUNX family of transcription factors and encodes a nuclear protein, a key transcription factor of osteoblast differentiation[42, 43]. A genetic variant in *RUNX2* (rs13191376) was found to be associated with CCT in a cross-ancestry GWAS [21]. Another genetic variant in *RUNX2* (rs1755056) was found to be significantly associated with IOP and weakly associated with POAG in a GWAS combing participants from the UK Biobank and published data from International Glaucoma Genetic Consortium[44]. In a transient transfection experiment using MG-63 cells and primary bovine corneal keratocytes, *RUNX2* transcription factors affected the expression of several small lecine rich proteoglycans (SLRP) including mimecan, biglycan and keratocan[45], which were shown to be important for the development and maintenance of corneal transparency[46-48]. In another study using the rabbit corneal epithelial cell line RCE1(5T5), RNAseq based transcriptome analysis showed *RUNX2* exhibited the highest expression in terminally differentiated corneal epithelial cells[49]. In breast cancer, it was found that *RUNX2* functioned through the androgen receptor to regulate prolactin-induced protein (PIP)[50], a new biomarker for keratoconus[51]. These findings suggested that *RUNX2* likely plays an important role in affecting CCT and the susceptibility of POAG and keratoconus.

Our study has some limitations. The number of probes used in our SMR analysis was limited, especially in the SMR analysis of the participants of East Asian ancestry. As a result, we may have missed some important genes. Moreover, the sample size and the number of eligible genetic variants for GWAS in the participants of East Asian ancestry is limited, compared to GWAS in the participants of European ancestry, which may affect the power of our SMR analysis and contribute the to insignificant findings in the participants of East Asian ancestry. The HEIDI test was significant for some of the identified genes (**Table 2**-**3**). As results, the possibility of horizontal pleiotropy, i.e., the identified association might be due to two distinct genetic variants in high linkage disequilibrium with each other, could not be ruled out. We only used eQTL data in the blood due to the unavailability of eQTL data from the eye. Our findings need to be validated in the future when eQTL data from the eye is available.

## Conclusions

We identified several genes pleiotropically associated with CCT, some of which represented novel genes influencing CCT. Our findings provided important leads to a better understanding of the genetic factors influencing CCT, and revealed potential therapeutic targets for the treatment of POAG and keratoconus.

## Data Availability

All data, models, or code generated or used during the study are available in a repository or online in accordance with funder data retention policies.The eQTL data can be downloaded at https://cnsgenomics.com/data/SMR/#eQTLsummarydata.The GWAS summarized data can be downloaded at https://datashare.is.ed.ac.uk/handle/10283/2976.

https://cnsgenomics.com/data/SMR/#eQTLsummarydata

https://datashare.is.ed.ac.uk/handle/10283/2976

## Acknowledgements

The study was supported by Beijing Tianjin Hebei Basic Research Cooperation Project (J200006), Pharmaceutical Collaborative Innovation Research Project of Beijing Science and Technology Commission (L192062), and National Key Research and Development Project (SQ2018YFC200148). Dr. Jingyun Yang’s research was supported by NIH/NIA grants P30AG10161, R01AG15819, R01AG17917, R01AG36042, U01AG61356 and 1RF1AG064312-01. Di Liu was supported by China Scholarship Council (CSC 201908110339).

## Author contribution

ZY, JY and WY designed and registered the study. DL and JY analyzed data and performed data interpretation. ZY, DL and JY wrote the initial draft and JY and WY contributed writing to subsequent versions of the manuscript. All authors reviewed the study findings and read and approved the final version before submission.

## Availability of data and materials

All data generated or analyzed during this study are included in this published article and its supplementary information files.

## Disclosure of Potential Conflicts of Interest

No potential conflicts of interest were disclosed by the authors.

## Notes

### Competing Interest Statement

The authors have declared no competing interest.

### Author Declarations

No IRB/oversight body was required for this study.

